# Can Artificial Intelligence Improve the Appropriate Use and Decrease the Misuse of REBOA?

**DOI:** 10.1101/2024.06.06.24308557

**Authors:** Mary Bokenkamp, Yu Ma, Ander Dorken-Gallastegi, Jefferson A. Proaño-Zamudio, Anthony Gebran, George C. Velmahos, Dimitris Bertsimas, Haytham M.A. Kaafarani, MPH

## Abstract

**BACKGROUND:** The use of resuscitative endovascular balloon occlusion of the aorta (REBOA) for control of noncompressible torso hemorrhage remains controversial. We aimed to utilize a novel and transparent/interpretable artificial intelligence (AI) method called Optimal Policy Trees (OPT), to improve the appropriate use and decrease the misuse of REBOA in hemodynamically unstable blunt trauma patients.

**METHODS:** We trained then validated OPTs that “prescribe” REBOA in a 50:50 split on all hemorrhagic shock blunt trauma patients in the 2010-2019 ACS-TQIP database based on rates of survival. Hemorrhagic shock was defined as a systolic blood pressure ≤ 90 on arrival or transfusion requirement of ≥ 4 units of blood in the first 4 hours of presentation. The expected 24-hour mortality rate following OPT prescription was compared to the observed 24-hour mortality rate in patients who were or were not treated with REBOA.

**RESULTS:** Out of 4.5 million patients, 100,615 were included and 803 underwent REBOA. REBOA patients had a higher rate of pelvic fracture, femur fracture, hemothorax, pneumothorax, and thoracic aorta injury (p<0.001). The 24-hour mortality rate for the REBOA vs. non-REBOA group was 47% vs. 21%, respectively (p<0.001). OPTs resulted in an 18% reduction in 24-hour mortality for REBOA and 0.8% reduction in non-REBOA patients.

**CONCLUSION:** Interpretable AI models can improve mortality in unstable blunt trauma patients by optimizing the use and decreasing the misuse of REBOA. These models to date have been used to predict outcomes, but their groundbreaking use will be prescribing interventions and changing outcomes.

**LEVEL OF EVIDENCE:** Level IV, Prognostic

## BACKGROUND

The use of resuscitative endovascular balloon occlusion of the aorta (REBOA) in trauma continues to be highly controversial. Its use was born during the Korean war as a temporary solution to control noncompressible torso hemorrhage in military personnel.^1^ While the use of REBOA in civilian trauma could be similarly beneficial, studies have not shown consistent results,^2-9^ and randomized control trials for its use have been difficult to design, view the real emergent nature of the situation warranting its use.^10-12^ Of the retrospective and prospective studies available, some have shown increased mortality^2-3^ while others claiming decreased mortality.^4-6^ Similarly, the studies vary in the reporting of the procedure-associated complications.^2-9^ As such, the indication and contraindications of its use continue to be a hotly debated subject in almost each trauma surgery conference and journal.

In recent years, our surgical-engineering collaborative group has successfully applied the power of Artificial Intelligence (AI) methodologies to surgical patients. We have previously leveraged a novel and interpretable AI technique called Optimal Classification Trees (OCT) to predict risk and outcomes in emergency general surgery^13^ and trauma patients.^14^ This resulted in two accurate and transparent algorithms that were translated into interpretable and user-friendly smartphone applications. These applications have since been downloaded and are in use by thousands of surgeons worldwide. For the present study, we aspired to utilize a different AI methodology developed at the Massachusetts Institute of Technology (MIT) called Optimal Policy Trees (OPT).^15-16^ Unlike OCTs which are “predictive”, OPTs are “prescriptive”. These trees work to “prescribe” the best treatment for different patient subgroups to achieve the best possible outcome of interest. The aim of this study was to utilize OPTs to improve the appropriate use and decrease the misuse of REBOA in hemodynamically unstable blunt trauma patients.

## METHODS

### Patient Population

The American College of Surgeons Trauma Quality Improvement Program (ACS-TQIP) Participant Use Data Files (PUFs) years 2010-2019 were used as the data source. Patients age > 16 who suffered a blunt traumatic injury and arrived at the hospital in hemorrhagic shock were included. Hemorrhagic shock was defined as a systolic blood pressure (SBP) ≤ 90 or a transfusion requirement of ≥ 4 units of red blood cells (RBCs) within the first 4 hours of hospital arrival. Patients were excluded if they were transferred in the first 24 hours or were missing data for age or length of stay (LOS). Patients who underwent REBOA placement within 4 hours of hospital arrival were identified using ICD-10 procedure codes (04L03DZ, 04L03DJ, 04L04DZ, 02LW3DJ, 04L04ZZ).

### Data Points

Multiple data points were collected. To ensure the OPTs would prescribe REBOA in a clinically meaningful way, the only data points that were utilized in creation of the trees were those that would be obtainable in the trauma bay. For example, at the point of deciding to use REBOA or not, the surgeon would know the patient’s approximate age and initial vitals but would not know the volume of blood products the patient would receive nor their ISS. These independent variables utilized were age, sex, admission physiology (systolic blood pressure (SBP), pulse, temperature, Glasgow Coma Scale (GCS), respiratory rate, pulse oximetry), use of supplemental oxygen, intubation in the emergency department (ED), height, weight, body mass index (BMI), signs of life upon emergency department (ED) arrival, hospital teaching status, and ACS verification level. Major injuries that could be suspected or diagnosed during the primary and secondary survey or initial radiological workup in the trauma bay were also included. These were pelvic fracture, femur fracture, hemothorax, pneumothorax, and thoracic aorta injury. Along a similar line of thinking, we included procedures that the patient may undergo in the trauma bay. These were chest tube placement, transfusion of RBC, and transfusion of whole blood. A time constraint of one hour was placed on these to capture procedures that were performed immediately upon arrival. The ED diagnoses and procedures were identified using ICD-10 codes (**Tables S1 and S2**).

Additional patient- and injury-related data points were analyzed separately to provide a more comprehensive picture of the overall cohort. The patients were stratified into two groups for comparison: those who underwent REBOA within 4 hours of hospital arrival (REBOA) and those that did not (No REBOA). The data points analyzed included blood products transfused within 4 hours of hospital arrival (red blood cells (RBC), fresh frozen plasma (FFP), platelets (PLT), and cryoprecipitate (Cryo), AIS, ISS, hemorrhage control procedures required within 4 hours of hospital arrival (laparotomy, thoracotomy/sternotomy, extremity vascular, preperitoneal pelvic packing (PPP), external fixation (EF) of pelvis, angioembolization (AE) of pelvis), and comorbidities. Laparotomy, thoracotomy/sternotomy, angioembolization, and extremity vascular are procedures tracked by ACS-TQIP. PPP and EF were identified using ICD-10 procedure codes (**Table S2**).

### Outcomes

The primary outcome targeted by the OPT model for improvement was 24-hour mortality. Secondary outcomes, analyzed separately, included hospital complications (catheter-associated urinary tract infection (CAUTI), central line-associated blood stream infection (CLABSI), superficial surgical site infection (SSI), deep SSI, organ space SSI, sepsis, pressure ulcer, deep vein thrombosis (DVT), pulmonary embolism (PE), compartment syndrome, cardiac arrest, myocardial infarction (MI), acute respiratory distress syndrome (ARDS), ventilator-associated pneumonia (VAP), acute kidney injury (AKI), and unplanned intubation, return to OR, or ICU admission), ventilator days, ICU LOS, and in-hospital mortality.

### Optimal Policy Trees

To create our AI-based prescriptive models, we used an innovative and interpretable machine-learning methodology called OPT.^15-16^ OPT leverages the power of mixed-integer programming by formulating the policy prescription problem as an optimization problem. The OPT method has been shown to achieve superior solutions to traditional machine-learning single-tree methods such as Classification and Regression Trees (CART) on various real-world datasets. Unlike many other “black box” machine-learning algorithms such as Regress and Compare or Causal Forest, OPT can make informed and accurate decisions without sacrificing intuitive understanding of how each decision is made. The ability to follow a transparent, concise decision path allows clinicians to understand exactly how each factor is incorporated into the decision-making process.

Through OPT, we produced a prescriptive model to improve 24-hour mortality in blunt hemodynamically unstable trauma patients who did or did not undergo REBOA placement. There were essentially five steps to accomplish this. First, the study population and data points as described above were isolated. Second, missing values for independent variables were imputed using a machine-learning method called Optimal Impute.^17^ Third, reward estimation was performed. This step estimates the probability that a given observation (patient) is assigned a given treatment (REBOA vs. No REBOA) and the probability of outcome (24-hr mortality) for each observation under each treatment option. Random forest classifiers are used to make these estimations. Doubly robust estimation is then used to construct reward matrices. These rewards are used to train an OPT to prescribe new treatments in such a way that the risk 24-hour mortality is reduced in comparison with the current risk of 24-hour mortality.

The fourth step involved model training and evaluation. Models are trained on a training and testing split of 50:50 to ensure sufficient data is saved to achieve high-quality reward estimation on the test set. Grid search is applied to select the best combination of hyperparameters (i.e. minimum number of samples in leaf, maximum depth of tree, complexity parameter) such that the best reward minimization is achieved in the training set. To avoid any information from the training set leaking through to the out-of-sample evaluation, instead of directly using the rewards from our existing reward estimator trained on the training set, we estimate a new set of reward estimators using only the test set and evaluate the policy against these rewards. Finally, the best performing tree was evaluated for clinical integrity and logic. If any inconsistencies were noted (for example, accidental inclusion of independent variables that would not be known at the time of REBOA placement), the data set was adjusted, and these steps were repeated until the final tree was obtained.

### Measurement of Model Performance

All propensity and outcome estimations are evaluated using the area under the receiver operating characteristic curve (ROC AUC). The AUC measures the ability of a model to discriminate between the outcomes of interest (24-hour mortality) and has been used extensively for binary classification problems due to its superior ability to account for problems such as class imbalance.

### Policy Evaluation

The expected 24-hour mortality rate following OPT prescription was compared to the observed 24-hour mortality rate in patients who were or were not treated with REBOA. This was done by calculating the average predicted probability of mortality under the treatments prescribed by the tree for the test set compared to the average probability of mortality under the treatment assignments that were observed. These results are reported as the absolute risk reduction (ARR). This was also calculated for each terminal leaflet of the tree to identify where the largest benefits were achieved by the prescriptive model.

### Other Statistical Analysis

The REBOA and No REBOA patient groups were compared using descriptive statistics. Categorical variables were compared using Pearson’s chi-squared test and continuous variables with the Kruskal-Wallis test. Categorical variables were reported as number of patients (percentage) and continuous variables were reported as median (interquartile range [IQR]). The level of significance was set at a p-value of < 0.05. All analyses were performed using STATA v.17 (StataCorp 2021, College Station, TX).

### Ethical Oversight

This study was submitted to and deemed exempt from approval by the Mass General Brigham Institutional Review Board.

## RESULTS

Out of a total of 4.5 million patients, 121,465 suffered blunt trauma and arrived to the hospital in hemorrhagic shock. After applying the aforementioned exclusion criteria, 100,615 patients comprised the study population. Within this group, 803 (0.8%) underwent REBOA within 4 hours of hospital arrival and 99,812 (99.2%) did not.

### Baseline Characteristics

The characteristics of REBOA vs. No REBOA patients utilized in creation of OPTs are displayed in **Table 1**. These are all data points that would have been theoretically known or obtainable during the primary and secondary survey. In summary, the REBOA patients were younger with a median age of 48 (IQR=29, 61, p<0.001) and mostly male (69.1%). They had a slightly higher SBP and pulse on arrival. The median GCS of REBOA patients was 3 (IQR=3, 14) compared to 14 (IQR=3, 15) for No REBOA patients. The majority of REBOA placements occurred at ACS Level 1 facilities (86.7%). Regarding ED procedures, 64.6% of REBOA patients underwent transfusion of RBCs vs. 19.9% of No REBOA patients. Of the ED diagnoses evaluated, REBOA patients had more injuries of all types with pneumothorax being most common (46.8%).

**Table 1.** Characteristics of REBOA versus No REBOA patients that were utilized in creation of OPTs.

|  | **No REBOA** | **REBOA** | **p-value** |
| --- | --- | --- | --- |
| **N (%)** | 99812 (99.2%) | 803 (0.8%) |  |
| **Age, median (IQR)** | 52 (33, 66) | 48 (29, 61) | <0.001 |
| **Sex, n (%)** |  |  | 0.008 |
| Male | 64471 (64.6%) | 555 (69.1%) |  |
| Female | 35317 (35.4%) | 248 (30.9%) |  |
| **Admission physiology, median (IQR)** |  |  |  |
| SBP | 82 (70, 90) | 86 (69, 118) | <0.001 |
| Pulse | 90 (70, 112) | 108 (80, 130) | <0.001 |
| Temperature | 36.4 (36, 36.7) | 36.1 (35.6, 36.5) | <0.001 |
| GCS | 14 (3, 15) | 3 (3, 14) | <0.001 |
| Respiratory rate | 18 (16, 22) | 20 (16, 25) | <0.001 |
| Pulse oximetry | 97 (93, 100) | 97 (91, 100) | 0.016 |
| **Supplemental oxygen, n (%)** | 49406 (55.4%) | 547 (79.3%) | <0.001 |
| **Intubated in ED, n (%)** | 25577 (25.6%) | 322 (40.1%) | <0.001 |
| **Height, median (IQR)** | 172.72 (165, 180) | 175 (165.1, 180.3) | 0.034 |
| **Weight, median (IQR)** | 80.29 (68, 97) | 85 (72, 100) | <0.001 |
| **BMI, median (IQR)** | 26.9 (23.5, 31.7) | 28.7 (24.7, 33.4) | <0.001 |
| **Signs of life in ED, n (%)** |  |  | 0.57 |
| No signs of life | 8628 (8.6%) | 74 (9.2%) |  |
| Signs of life | 91184 (91.4%) | 729 (90.8%) |  |
| **Teaching status, n (%)** |  |  | <0.001 |
| Community | 34933 (35.1%) | 187 (23.4%) |  |
| Nonteaching | 11753 (11.8%) | 52 (6.5%) |  |
| University | 52735 (53.0%) | 559 (70.1%) |  |
| **ACS verification level, n (%)** |  |  | <0.001 |
| 1 | 49277 (58.8%) | 589 (86.7%) |  |
| 2 | 21719 (25.9%) | 80 (11.8%) |  |
| 3 | 12848 (15.3%) | 10 (1.5%) |  |
| **ED Procedures, n (%)** |  |  |  |
| Chest tube placement | 942 (0.9%) | 24 (3.0%) | <0.001 |
| Transfusion RBCs | 19865 (19.9%) | 519 (64.6%) | <0.001 |
| Transfusion whole blood | 1643 (1.6%) | 77 (9.6%) | <0.001 |
| **ED Diagnoses, n (%)** |  |  |  |
| Pelvic fx | 9105 (9.1%) | 287 (35.7%) | <0.001 |
| Femur fx | 2077 (2.1%) | 33 (4.1%) | <0.001 |
| Hemothorax | 6851 (6.9%) | 160 (19.9%) | <0.001 |
| Pneumothorax | 20938 (21.0%) | 376 (46.8%) | <0.001 |
| Thoracic aorta injury | 2419 (2.4%) | 51 (6.4%) | <0.001 |

Abbreviations: SBP: systolic blood pressure, GCS: Glasgow Coma Scale, ED: emergency department, BMI: body mass index, ACS: American College of Surgeons, RBC: red blood cell, Fx: fracture

The other injury-related characteristics of REBOA vs. No REBOA patients that were not included in creation of OPTs are displayed in **Table 2**. REBOA patients received more units of all blood products (RBC, FFP, PLT, Cryo). The median ISS for REBOA vs. No REBOA patients was 34 (IQR=26, 45) and 21 (IQR=10, 33), respectively. The highest AIS body region score for REBOA patients were thorax, abdomen, and extremity. The most common hemorrhage control procedure performed on REBOA patients within 4 hours of hospital arrival was laparotomy (46.5%) followed by pelvic angioembolization (15.2%).

**Table 2.**
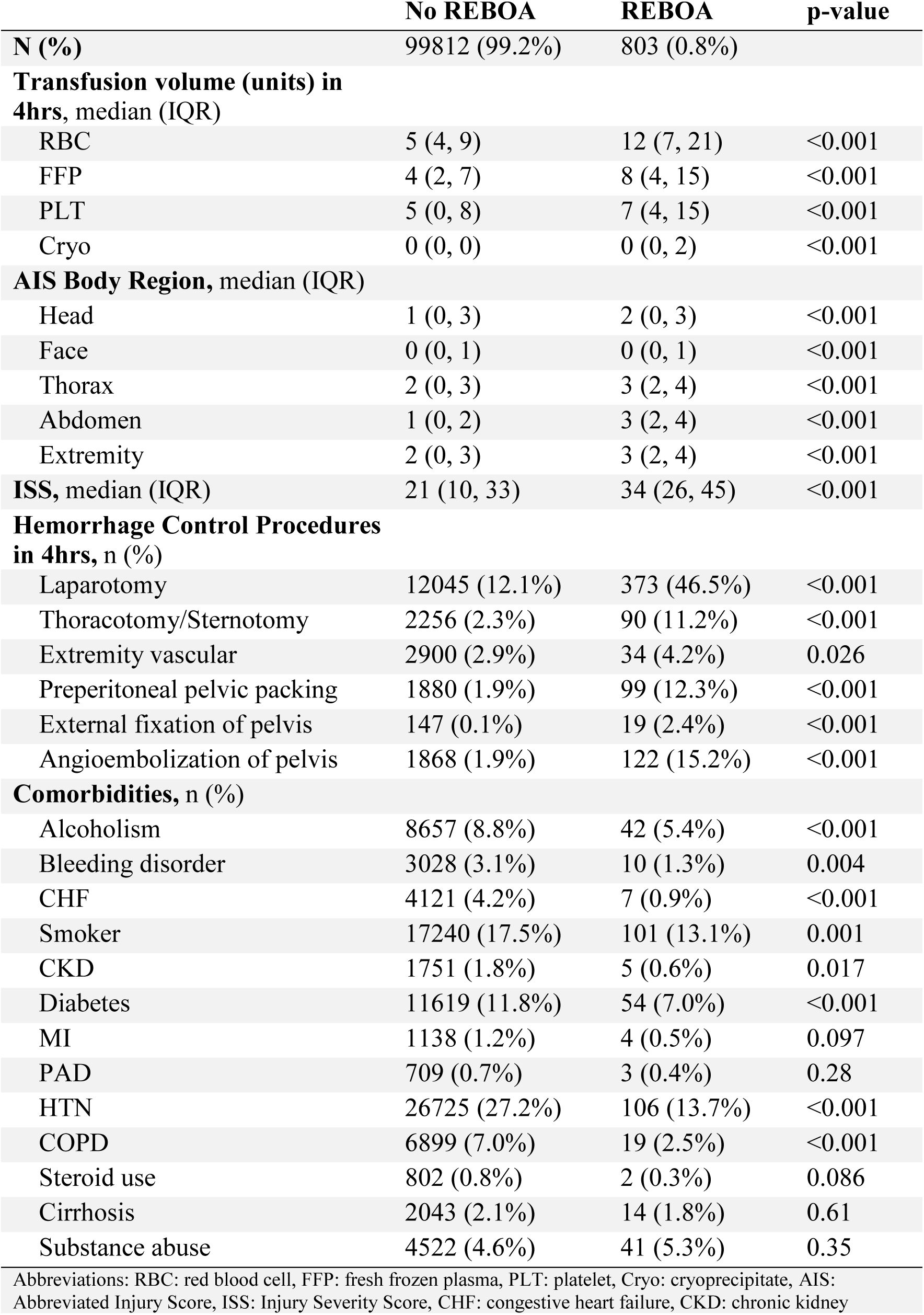
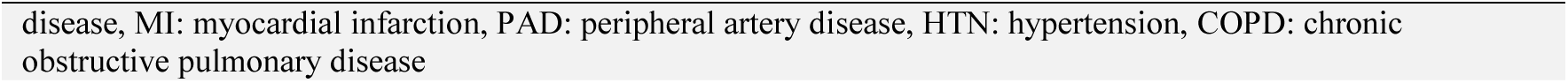
Other injury-related characteristics of REBOA versus No REBOA patients that were not included in creation of OPTs.

|  | No REBOA | REBOA | p-value |
| --- | --- | --- | --- |
| **N (%)** | 99812 (99.2%) | 803 (0.8%) |  |
| **Transfusion volume (units) in 4hrs, median (IQR)** |  |  |  |
| RBC | 5 (4, 9) | 12 (7, 21) | <0.001 |
| FFP | 4 (2, 7) | 8 (4, 15) | <0.001 |
| PLT | 5 (0, 8) | 7 (4, 15) | <0.001 |
| Cryo | 0 (0, 0) | 0 (0, 2) | <0.001 |
| **AIS Body Region, median (IQR)** |  |  |  |
| Head | 1 (0, 3) | 2 (0, 3) | <0.001 |
| Face | 0 (0, 1) | 0 (0, 1) | <0.001 |
| Thorax | 2 (0, 3) | 3 (2, 4) | <0.001 |
| Abdomen | 1 (0, 2) | 3 (2, 4) | <0.001 |
| Extremity | 2 (0, 3) | 3 (2, 4) | <0.001 |
| **ISS, median (IQR)** | 21 (10, 33) | 34 (26, 45) | <0.001 |
| **Hemorrhage Control Procedures in 4hrs, n (%)** |  |  |  |
| Laparotomy | 12045 (12.1%) | 373 (46.5%) | <0.001 |
| Thoracotomy/Sternotomy | 2256 (2.3%) | 90 (11.2%) | <0.001 |
| Extremity vascular | 2900 (2.9%) | 34 (4.2%) | 0.026 |
| Preperitoneal pelvic packing | 1880 (1.9%) | 99 (12.3%) | <0.001 |
| External fixation of pelvis | 147 (0.1%) | 19 (2.4%) | <0.001 |
| Angioembolization of pelvis | 1868 (1.9%) | 122 (15.2%) | <0.001 |
| **Comorbidities, n (%)** |  |  |  |
| Alcoholism | 8657 (8.8%) | 42 (5.4%) | <0.001 |
| Bleeding disorder | 3028 (3.1%) | 10 (1.3%) | 0.004 |
| CHF | 4121 (4.2%) | 7 (0.9%) | <0.001 |
| Smoker | 17240 (17.5%) | 101 (13.1%) | 0.001 |
| CKD | 1751 (1.8%) | 5 (0.6%) | 0.017 |
| Diabetes | 11619 (11.8%) | 54 (7.0%) | <0.001 |
| MI | 1138 (1.2%) | 4 (0.5%) | 0.097 |
| PAD | 709 (0.7%) | 3 (0.4%) | 0.28 |
| HTN | 26725 (27.2%) | 106 (13.7%) | <0.001 |
| COPD | 6899 (7.0%) | 19 (2.5%) | <0.001 |
| Steroid use | 802 (0.8%) | 2 (0.3%) | 0.086 |
| Cirrhosis | 2043 (2.1%) | 14 (1.8%) | 0.61 |
| Substance abuse | 4522 (4.6%) | 41 (5.3%) | 0.35 |

Abbreviations: RBC: red blood cell, FFP: fresh frozen plasma, PLT: platelet, Cryo: cryoprecipitate, AIS: Abbreviated Injury Score, ISS: Injury Severity Score, CHF: congestive heart failure, CKD: chronic kidney
disease, MI: myocardial infarction, PAD: peripheral artery disease, HTN: hypertension, COPD: chronic obstructive pulmonary disease

### Outcomes

The primary and secondary outcomes analyzed are reported in **Table 3**. In summary, REBOA patients had a 46.9% 24-hour mortality rate vs. 20.9% in No REBOA patients (p<0.001, **Table 3**). This rate was the target of the OPT model which is discussed in the next section. The in-hospital mortality rates for REBOA vs. No REBOA patients were 61.8% and 30.8%, respectively (p<0.001). The rates of several complications such as CAUTI, deep SSI, sepsis, pressure ulcer, DVT, PE, compartment syndrome, cardiac arrest, AKI, and unplanned return to OR were statistically higher. In addition, REBOA patients had more ventilator and ICU days.

**Table 3.**
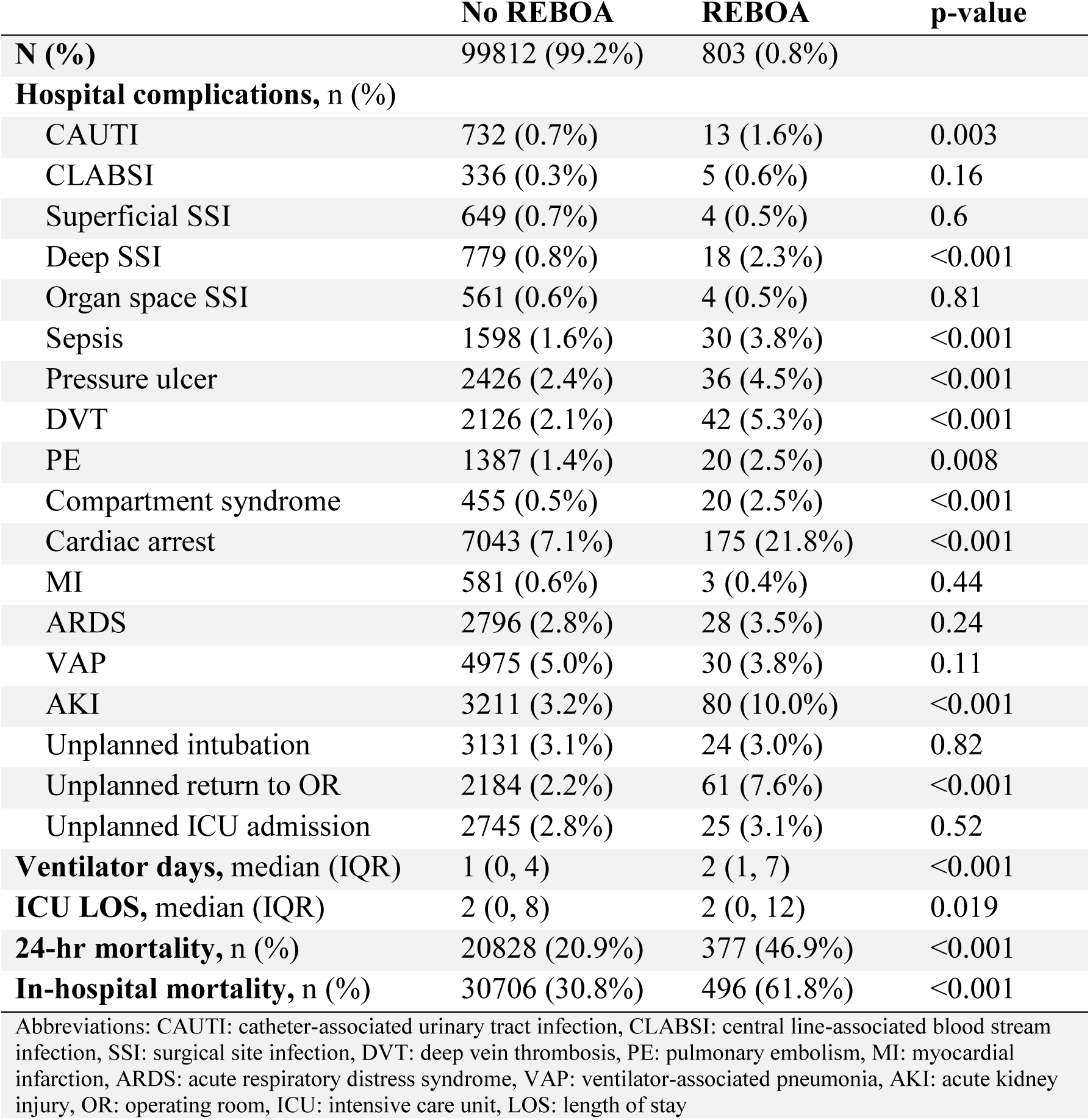
Outcomes of REBOA versus No REBOA patients.

|  | **No REBOA** | **REBOA** | **p-value** |
| --- | --- | --- | --- |
| **N (%)** | 99812 (99.2%) | 803 (0.8%) |  |
| **Hospital complications, n (%)** |  |  |  |
| CAUTI | 732 (0.7%) | 13 (1.6%) | 0.003 |
| CLABSI | 336 (0.3%) | 5 (0.6%) | 0.16 |
| Superficial SSI | 649 (0.7%) | 4 (0.5%) | 0.6 |
| Deep SSI | 779 (0.8%) | 18 (2.3%) | <0.001 |
| Organ space SSI | 561 (0.6%) | 4 (0.5%) | 0.81 |
| Sepsis | 1598 (1.6%) | 30 (3.8%) | <0.001 |
| Pressure ulcer | 2426 (2.4%) | 36 (4.5%) | <0.001 |
| DVT | 2126 (2.1%) | 42 (5.3%) | <0.001 |
| PE | 1387 (1.4%) | 20 (2.5%) | 0.008 |
| Compartment syndrome | 455 (0.5%) | 20 (2.5%) | <0.001 |
| Cardiac arrest | 7043 (7.1%) | 175 (21.8%) | <0.001 |
| MI | 581 (0.6%) | 3 (0.4%) | 0.44 |
| ARDS | 2796 (2.8%) | 28 (3.5%) | 0.24 |
| VAP | 4975 (5.0%) | 30 (3.8%) | 0.11 |
| AKI | 3211 (3.2%) | 80 (10.0%) | <0.001 |
| Unplanned intubation | 3131 (3.1%) | 24 (3.0%) | 0.82 |
| Unplanned return to OR | 2184 (2.2%) | 61 (7.6%) | <0.001 |
| Unplanned ICU admission | 2745 (2.8%) | 25 (3.1%) | 0.52 |
| **Ventilator days, median (IQR)** | 1 (0, 4) | 2 (1, 7) | <0.001 |
| **ICU LOS, median (IQR)** | 2 (0, 8) | 2 (0, 12) | 0.019 |
| **24-hr mortality, n (%)** | 20828 (20.9%) | 377 (46.9%) | <0.001 |
| **In-hospital mortality, n (%)** | 30706 (30.8%) | 496 (61.8%) | <0.001 |

Abbreviations: CAUTI: catheter-associated urinary tract infection, CLABSI: central line-associated blood stream infection, SSI: surgical site infection, DVT: deep vein thrombosis, PE: pulmonary embolism, MI: myocardial infarction, ARDS: acute respiratory distress syndrome, VAP: ventilator-associated pneumonia, AKI: acute kidney injury, OR: operating room, ICU: intensive care unit, LOS: length of stay

### OPT and 24-hour Mortality

Figure 2 shows the OPT model for prescribing REBOA or No REBOA to blunt trauma patients in hemorrhagic shock to improve 24-hour mortality. The tree is transparent and interpretable with a relatively small number of decision branches. Each rectangular box represents a “leaf”, and the data point the model used at each as a branch point is listed below it. Within each leaf, either “Prescribe No REBOA” or “Prescribe REBOA” is the treatment the model prescribed. The “N” number of patients at each leaf are also reported. The color of the terminal leaflets represents each treatment and the prescription strength. The blue color corresponds to REBOA and red to No REBOA. If the color is very solid or dark, it means the prescription in that node is very confident. If the color is pale or light, it means that the difference between prescribing REBOA vs. No REBOA is less prominent.

**Figure 1.**
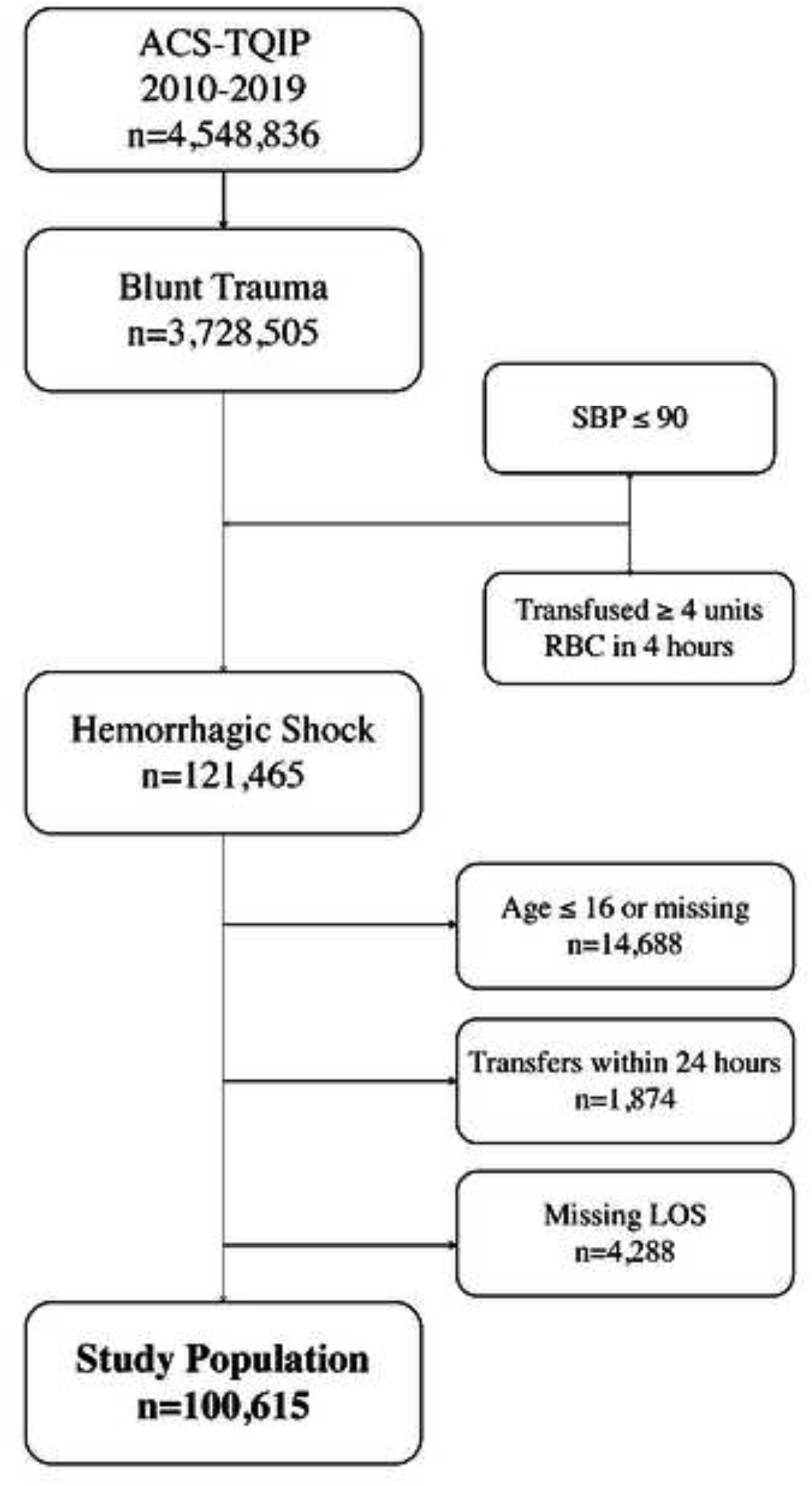
Flow chart for the study population.

**Figure 2.**
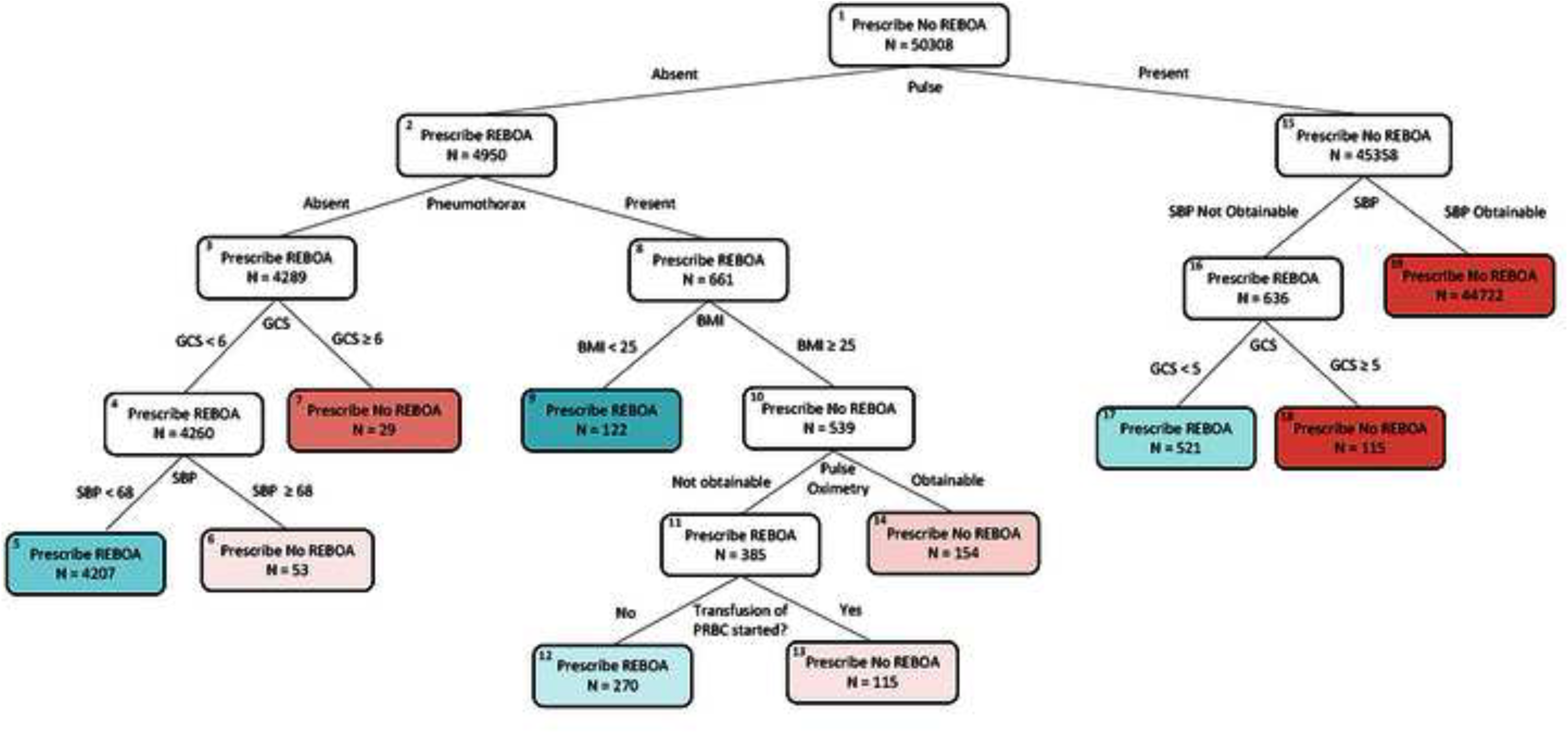
Optimal Policy Tree (OPT) that prescribes REBOA versus No REBOA to improve 24-hour mortality in hemodynamically unstable blunt trauma patients.

Starting at the top of the tree at Leaf #1, one can appreciate the “Prescribe No REBOA” and N=50,308, which represents the 50:50 training to testing split that was performed. The first branch point asks if the patient had a pulse or not. If present, the next Leaf #15 states “Prescribe No REBOA.” The model suggests these patients not receive REBOA yet, but to have a more confident prescription one needs to continue going through the tree. The next branch point is SBP. If SBP was obtainable, Leaf #19 prescribes No REBOA and is the terminal leaf. If SBP was not obtainable, the tree asks about GCS. If the GCS ≥ 5, the model prescribes No REBOA. If the GCS < 5, the tree prescribes REBOA.

Returning to Leaf #1, we can follow the model to the left for patients with no pulse. The next Leaf #2 uses pneumothorax as a branch point. If there is no pneumothorax, the model next asks about GCS. If GCS ≥ 6, the model prescribes No REBOA. If GCS < 6, the model prescribes REBOA. The next branch point uses SBP of 68. For patients with an obtainable SBP ≥ 68, the model prescribes No REBOA and for patients with SBP < 68, the model prescribes REBOA. The tree can be similarly followed through each leaf and branch point to the terminal leaflets.

The ARR in 24-hour mortality rate for the overall study population, REBOA patients, and No REBOA patients can be visualized in Figure 3. The ARR in the overall study population and No REBOA patients was about the same at 0.9% and 0.8%, respectively. The ARR was largest amongst REBOA patients, with the original cohort having a 47% 24-hour mortality rate and a prescribed 29% 24-hour mortality rate, resulting in an ARR of 18%. When the terminal leaflets were examined, the largest benefit was seen at leaflet #5 (ARR=7.98%) and #17 (ARR=5.44%) (**Table 4**).

**Figure 3.**
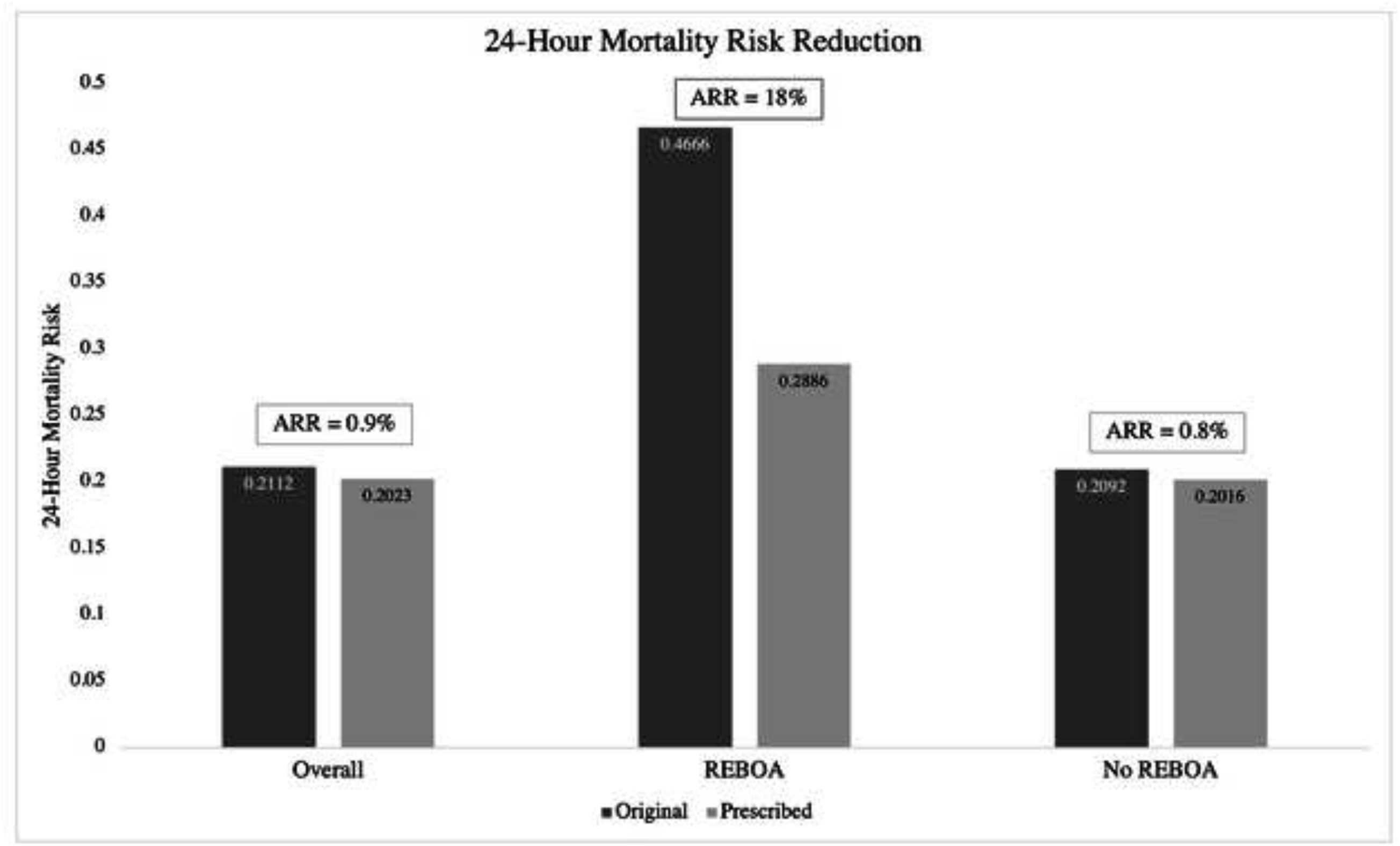
Absolute risk reduction (ARR) in 24-hour mortality for the overall study population, REBOA patients, and No REBOA patients.

**Table 4.** Terminal leaf level absolute risk reduction (ARR) of original vs. prescribed REBOA treatment on 24-hour mortality. Leaf numbers correspond to those in Figure 2.

| **Leaf #** | **Original** | **Prescribed** | **ARR** | **ARR (%)** |
| --- | --- | --- | --- | --- |
| **5** | 0.959 | 0.879 | 0.080 | 7.984 |
| **6** | 0.719 | 0.718 | 0.001 | 0.080 |
| **7** | 0.406 | 0.406 | 0.000 | 0.000 |
| **9** | 0.848 | 0.834 | 0.015 | 1.477 |
| **12** | 0.919 | 0.888 | 0.031 | 3.059 |
| **13** | 0.888 | 0.888 | 0.000 | 0.005 |
| **14** | 0.820 | 0.819 | 0.001 | 0.087 |
| **17** | 0.740 | 0.685 | 0.054 | 5.438 |
| **18** | 0.190 | 0.182 | 0.008 | 0.831 |
| **19** | 0.123 | 0.121 | 0.002 | 0.153 |

## DISCUSSION

Utilizing a novel, transparent, and interpretable AI methodology called OPT, we have thus created a proof of concept model for the prescription of REBOA in hemodynamically unstable blunt trauma patients that can potentially decrease the 24-hour mortality of this high-risk population. The OPT model resulted in an ARR in 24-hour mortality of 0.8% in No REBOA patients and 18% in REBOA patients. In other words, our model showed very minimal improvement in 24-hour mortality when prescribing REBOA to those who did not receive it in real life, and a large improvement in 24-hour mortality when prescribing No REBOA to patients who did. These results suggest REBOA is potentially being overused in this patient population.

This is the first study to our knowledge that employs AI to elucidate indications for REBOA. Furthermore, this is the first study to utilize OPTs for decision-making in trauma. We perceive this model as a prototype for the use of AI to assist surgeons in decision-making, rather than advocate that this decision tree be the new guide for REBOA use. Further studies with more granular data can lead to more powerful OPT tree and decision-making tool creation.

Over the past few decades, substantial effort has been made to study the use REBOA for aortic occlusion in hemorrhaging trauma patients.^1-12, 18-19^ Despite these endeavors, there are currently no universally agreed upon indications for use and mixed data on outcomes keep the debate alive. Although minimally invasive, REBOA has been suggested to have significant complications including common femoral artery injury, aortoiliac injury, balloon rupture, and sequalae from prolonged aortic occlusion such as spinal cord injury, AKI, and multisystem organ failure.^10-11^ A recent study by Moore et al. prospectively observed patients from six US Level 1 trauma centers and found balloon inflation to increase SBP and achievement of return of spontaneous circulation (ROSC) in more than 50% of patients in cardiac arrest.^18^ These centers used a new ER-REBOA catheter, supporting the notion that advances in technology may improve outcomes surrounding REBOA use.

While these new results are encouraging, one of the most concerning aspects of REBOA remains to be the discrepancy in mortality outcomes reported. Two well-known studies have shown increased rates of mortality.^2-3^ Other studies have shown improved survival in various trauma populations.^4-6^ In the updated joint statement from the American College of Surgeons Committee on Trauma, American College of Emergency Physicians, National Association of Emergency Medical Services Physicians, and National Association of Emergency Medical Technicians, the authors note that none of the current evidence has shown that REBOA improves outcomes or survival compared to the current standard of treatment.^11^ While some studies show promising improvements, and the technology and technique of REBOA continue to evolve, our results suggest that REBOA was overused in the last few years and perhaps highlight the lack of indications and variation of use among different trauma centers.

The concern of bias that can be accidentally built into predictive and prescriptive AI models is real.^20-21^ In this study, the difficulty was mainly the lack of additional, dynamic and nuanced data points available in the trauma bay but not in the database for decision-making. While we did our best to simulate the pieces of information that would be known, this was certainly a limitation. This is an explanation for why seemingly irrelevant variables were used by the model, such as BMI and pulse oximetry (Figure 2). The AI algorithms are only as adept as the data used to create and train them.^21^

Alternatively, it was also intriguing to note which variables the algorithm did and did not use as this shows how influential they were to alter the outcome of interest. While BMI and pulse oximetry seem inconsequential, there may an underlying significance we have yet to uncover as surgeon-scientists. Interestingly, the model did not select pelvic fracture, an injury that is often associated with REBOA use. It is also worth discussing the fact that the leaflets with the highest ARR were #5 and #17. Leaf #5 prescribes REBOA to patients who had an absent pulse, no pneumothorax, GCS < 6, and SBP < 68. Leaf #17 prescribes REBOA to patients who had a pulse, no obtainable SBP, and GCS < 5. Our model aligns with the general indications that have been used for REBOA to date: hypotensive blunt trauma patients.

There are several other limitations to this study. First, the AI model was constructed using retrospective data from a large national databank that was not designed with REBOA in mind. There is no data on what model of REBOA was used, introducer sheath size, time to access, insertion, and inflation and associated SBP at those times, level of aortic occlusion, or duration of aortic occlusion. Second, ACS-TQIP also has no data on the FAST exam, which is a critical missing piece of information for REBOA use. Third, there are only a small number of high-volume trauma centers that use REBOA, and this could have a clustering effect in the data. Finally, we would like to reiterate our intention that this be appreciated as proof of the conceptual idea behind prescriptive AI methodologies and its use in trauma patients. The key to improving these models will continue to be the application of an appropriate, robust data set.

## CONCLUSION

Our study is a proof of concept one to utilize the AI non-linear logic to improve the use and decrease the misuse of REBOA. Our algorithms suggest that REBOA may have been overused in blunt hemodynamically unstable trauma patients in the last few years, and improvement of the decision making with the assistance of AI can potentially result in 18% ARR in 24-hour mortality for patients by avoiding the use of REBOA. Our model is not ready for bedside use, and further studies with more granular data can improve its performance further for clinical practice. However, our study shows the premise that interpretable AI models can in the future improve mortality in unstable blunt trauma patients by optimizing decision-making and assisting surgeons in improving outcomes.

## Data Availability

All data produced are available online at The American College of Surgeons Trauma Quality Improvement Program (ACS-TQIP) Participant Use Data Files (PUFs).

https://www.facs.org/quality-programs/trauma/quality/tqp-participant-hub/

